# Association of urinary peptides with hypertension

**DOI:** 10.1101/2023.12.14.23299991

**Authors:** Emmanouil Mavrogeorgis, Margarita Kondyli, Harald Mischak, Antonia Vlahou, Justyna Siwy, Peter Rossing, Archie Campbell, Carina MC Mels, Christian Delles, Jan A Staessen, Agnieszka Latosinska, Alexandre Persu

**Affiliations:** Mosaiques Diagnostics GmbH, Hannover, Germany; Institute for Molecular Cardiovascular Research (IMCAR), RWTH Aachen University Hospital, Aachen, Germany; University of Patras, Department of Pharmacy, Patras, Greece; Center of Systems Biology, Biomedical Research Foundation of the Academy of Athens, Athens, Greece; Steno Diabetes Center Copenhagen, Herlev, Denmark; Department of Clinical Medicine, University of Copenhagen, Copenhagen, Denmark; Centre for Genomic and Experimental Medicine, Institute of Genetics and Cancer, University of Edinburgh, Edinburgh, UK; Hypertension in Africa Research Team (HART), Faculty of Health Sciences, North-West University, Potchefstroom, South Africa; MRC Research Unit for Hypertension and Cardiovascular Disease, North-West University, Potchefstroom, South Africa; School of Cardiovascular and Metabolic Health, University of Glasgow, Glasgow, UK; Research Institute Alliance for the Promotion of Preventive Medicine, Mechelen, Belgium; Division of Cardiology, Department of Cardiovascular Diseases, Cliniques Universitaires Saint-Luc and Pole of Cardiovascular Research, Institut de Recherche Expérimentale et Clinique, Université Catholique de Louvain, Brussels, Belgium

**Keywords:** blood pressure, CE-MS, hypertension, peptides, urine

## Abstract

**BACKGROUND:** Hypertension is a common condition worldwide, yet its underlying mechanisms remain largely unknown. This study aims at identifying urinary peptides associated with hypertension to further explore its molecular pathophysiology.

**METHODS:** Peptidome data from 2876 individuals without end-organ damage were retrieved from the Human Urinary Proteome Database general population (discovery) or type 2 diabetic (validation) cohorts. Participants were divided based on systolic and diastolic blood pressure (SBP and DBP) into hypertensive (SBP≥140mmHg and/or DBP≥90mmHg) and normotensive (SBP<120mmHg and DBP<80mmHg, without antihypertensive treatment) groups. Differences in peptide abundance between the two groups were confirmed using an external cohort (n=420) of participants without end-organ damage, matched for age, body-mass index, eGFR, sex and presence of diabetes. Further, associations of the peptides with BP as a continuous variable were investigated. Findings were compared with peptide biomarkers of chronic diseases and bioinformatics analyses were conducted to potentially highlight the underlying molecular mechanisms.

**RESULTS:** Between hypertensive and normotensive individuals, ninety-six (mostly COL1A1 and COL3A1) peptides were found significantly different in the discovery (adjusted) as well as the validation (nominal significance) cohorts with consistent regulation. Of these peptides, 83 were also consistently regulated in the matched cohort. A weak, yet significant association between their abundance and standardized BP was also observed.

**CONCLUSIONS:** Hypertension is associated with an altered urinary peptide profile, with evident collagen differential regulation. Peptides related to vascular calcification and sodium regulation are also affected. Whether these modifications reflect the pathophysiology of hypertension *per se* and/or early subclinical target organ damage warrants further investigation.

**Novelty and Relevance:** *What is New?:* This is the first study demonstrating differential regulation of urinary peptides in hypertensive patients, independent from other co-factors like age, diabetes, or established kidney or cardiovascular disease.

*What is Relevant?:* The observed changes in urinary peptides indicate individual differences in molecular changes observed in hypertension, and may guide personalized treatment based on the observed molecular changes

*Clinical/Pathophysiological Implications?:* The results indicate that collagen homeostasis may be a key molecular feature in hypertension and may serve as an attractive mechanism for pharmacological intervention.

## Background

Hypertension, a common condition referring to elevated blood pressure (BP), affects billions of adults aged 30-79 years worldwide, the majority of which are based in countries of low- and middle-income ^1^. Although hypertension is associated, among others, with heart disease, stroke, kidney damage and premature deaths, 80% of the hypertensive patients do not receive appropriate treatment^2^. The American Heart Association defines hypertension as BP≥130/80 mmHg, whereas European guidelines as BP≥140/90 mmHg. These different definitions, as expected, correspond to different prevalence, specifically 43.5% versus 18.9%, respectively^3^. Despite the numerous available treatments and lifestyle adaptations to lower BP, an estimated ∼10 million US adults show resistance to treatment^4,5^, indicating that the condition still requires improved and/or new multimodal therapeutic approaches.

BP is a highly complex and heterogeneous trait, to which several different causal factors might contribute ^6^ and the exact biological underlying mechanisms remain largely unknown. A complex interplay of various physiological mechanisms involving the kidneys, various hormonal systems, the autonomic nervous system and baroreceptors coordinate the regulation of BP^7^. Although these mechanisms ensure that BP is maintained within a narrow range for proper tissue perfusion, 95% of hypertensive patients are diagnosed with primary hypertension, occurring in the absence of secondary causes^6^.

This complex and mechanistically largely unexplored condition also appears to be interconnected with several chronic diseases. Along these lines, hypertension appears to have a crucial role not only as a major risk factor, but also as a pathophysiological consequence that further exacerbates chronic disease. In previous studies, focusing on chronic diseases (heart failure^8^, coronary artery^9^ and chronic kidney disease^10^, among others^11^), we determined differential regulation in the abundance of hundreds of specific urinary peptides, likely reflecting disease pathophysiology.

In an effort to contribute to the deciphering of the molecular mechanisms involved in hypertension, we aimed here to identify urinary peptides that differ significantly in patients with hypertension compared to normotensive individuals. To this end, the urinary peptide data of almost 3000 participants of either population-based or diabetic cohort studies were obtained from the Human Urinary Proteome Database ^12^ and served as the basis for a series of statistical and bioinformatics analyses to further explore the molecular players associated with this trait.

## Methods

Complete, fully anonymized clinical and peptide data can be received upon reasonable request by contacting the corresponding author.

### Study Population

Considered were anonymized data from the FLEMENGHO^13^ and Generation Scotland: Scottish Family Health Study^8,14^ (general population) as well as DIRECT2^15^ and PRIORITY^16,17^ (diabetes type 2) since these studies were the largest of their kind in the database. On this initial extracted cohort of n=4726, the following exclusion criteria were applied: established end-organ damage, including estimated glomerular filtration rate (eGFR)<60 ml/min/1.73m^2^ (CKD-EPI), macroalbuminuria (total excretion of urinary albumin>300 mg/24h), left ventricular hypertrophy, heart failure, ischemic heart disease/CAD, transplantation, history of cancer, history of kidney or cardiovascular disease. The general population cohort was investigated for discovery purposes (n=749), while the diabetic cohort served for validation (n=2127). The SBP and DBP values were used to divide each cohort into two groups based on the European guidelines’ definition for hypertension, specifically, SBP≥140 mmHg and/or DBP≥90 mmHg (hypertensive), as well as SBP<120 mmHg and DBP<80 mmHg and not undergoing anti-hypertensive treatment (normotensive). An additional validation cohort of hypertensive and normotensive individuals (n=1352) matched for age, body-mass index (BMI), eGFR, sex and presence of diabetes was studied in an attempt to further confirm the results and findings. Inclusion and exclusion criteria were similar to the aforementioned discovery and validation cohorts.

### Urinary peptidomics

The data were extracted from the Human Urinary Proteome Database^12^, containing datasets acquired using the capillary electrophoresis coupled to mass spectrometry technique as described in several sources ^12,18–20^. Data were evaluated using the MosaFinder software and normalized based on the abundance of 29 collagen peptides insensitive to disease state^21^. Of the 5071 peptides identified to date, only those being present in at least 50% of the entire (discovery+validation) cohort of 2876 individuals, i.e., 888 peptides, remained for further analyses. Before any statistical analyses, the missing values of that dataset per column were replaced by randomly generated deviates of a uniform distribution in the range between 0.75*x and 1.25*x (where x is the column minimum value), similar to the approach described^22^. A detailed scheme of the study design and workflow is depicted in **Figure 1**.

**Figure 1.**
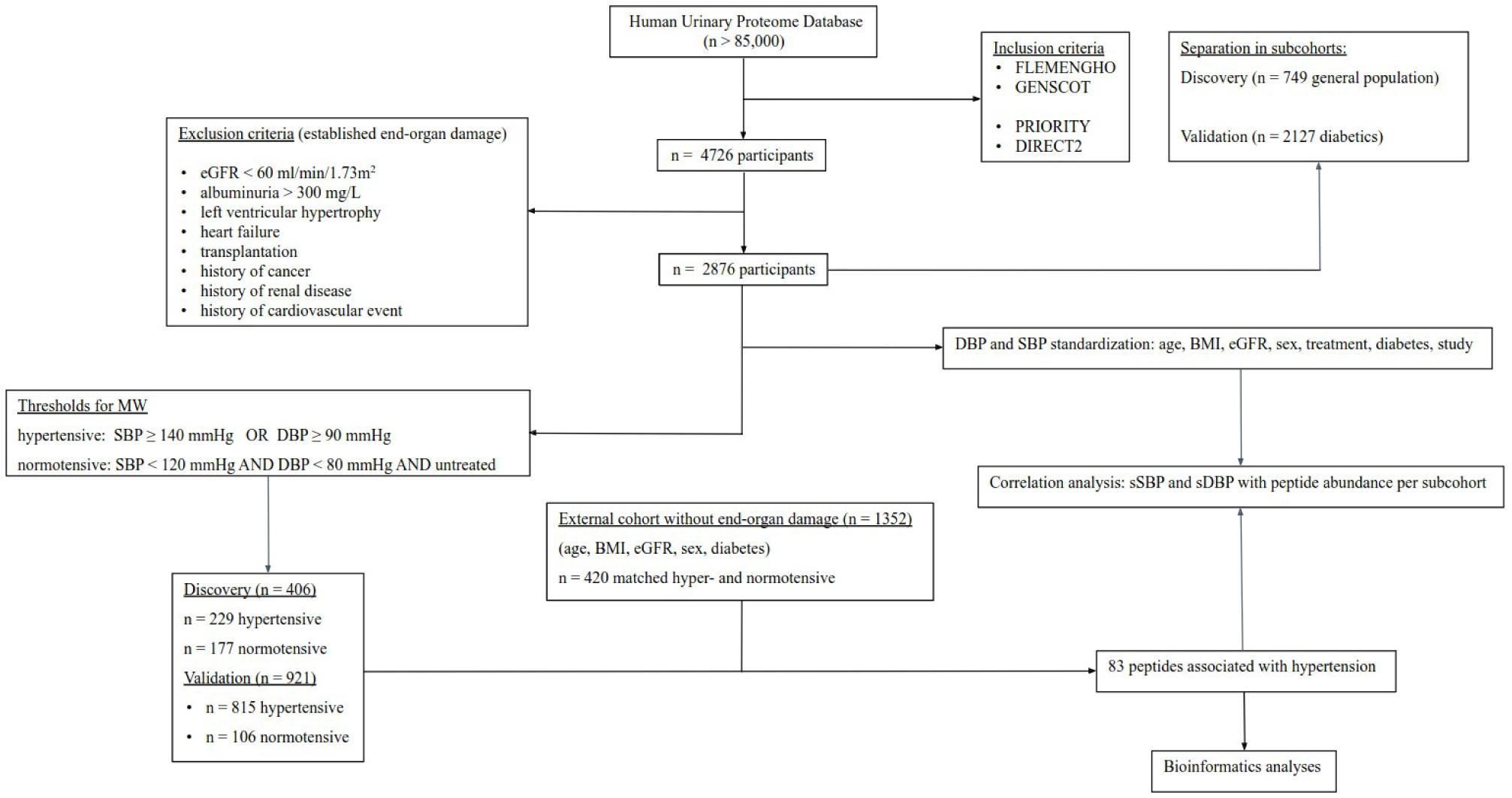
Study design and workflow. Study design. The urinary peptide data of 4726 participants belonging to either general population or type 2 diabetes studies were acquired from the Human Urinary Proteome Database^12^. After exclusion criteria related to established end-organ damage were applied, 2876 participants were considered for further analyses. Then, the individuals were assigned into discovery (general population) and validation sets (diabetes) and within each cohort participants were defined as hypertensive (SBP≥140mmHg and/or DBP≥90mmHg) and normotensive (SBP<120mmHg and DBP<80mmHg, without antihypertensive treatment), according to the current European guidelines. Individuals not matching either criteria were excluded. Eighty-three peptides demonstrating significantly different levels between the groups in each cohort (adjusted and nominal significance in the discovery and validation sets, respectively) as well as consistent regulation in these two cohorts and an external, matched cohort of n=420 participants (again, without established end-organ damage), appeared to be associated with hypertension. These peptides were subsequently used as an input for a correlation analysis based on the entire cohort (n=2876) between the peptide abundance and the standardized (for disease confounders, namely, age, BMI, eGFR, sex, presence of treatment, presence of diabetes as well as the specific cohort per individual) BP measures, in order to highlight the relevant associations. Last, bioinformatics analyses revealed the relevant peptide biological setting. BP: blood pressure. sSBP: standardized systolic blood pressure. sDBP: standardized diastolic blood pressure. BMI: body mass index. eGFR: estimated glomerular filtration rate. BH: Benjamini-Hochberg.

### Statistical analysis

#### Differential peptide abundance between the groups

Initially, a comparison of the peptide abundances between the hypertensive and normotensive groups was performed in the discovery cohort. Next, the significantly different peptides (applying Benjamini-Hochberg correction for multiple testing) were assessed in terms of consistent regulation and nominal significance in the validation cohort. Peptides found significantly affected were evaluated based on their consistency in regulation using the external matched cohort before being considered for further analyses, to account for the potential impact of confounders (age, BMI, eGFR, sex and presence of diabetes). Linear regression was used to examine a potential association of the hypertension-associated peptides with the respective significant changes of previously identified biomarkers in chronic diseases, namely, heart failure ^8^, coronary artery disease^9^ and chronic kidney disease^10^. The differential peptide abundance analysis was performed using the Mann-Whitney U test. A significance threshold of p-value<0.05 was applied, unless stated differently.

### Blood pressure standardization and correlation with peptide abundance

The following clinical parameters were considered as variables for the BP standardization: age, BMI, eGFR, sex, presence of antihypertensive treatment, presence of diabetes and study cohort. These variables were used as a basis for a stepwise linear regression applying the following approach (also described in ^22^): A specific p-value threshold was required for a variable to enter (p-value<0.15) or be removed (p-value>0.15) from the model. Initially a model was developed relying only on the variable with the lowest p-value. Subsequently, from the variables not included yet in the model, the one with the lowest p-value was further added to the previous model. Then, the variable already present in the model (before the addition of that second, newly added variable), was checked again in terms of p-value and was removed from the model if it exceeded the designated threshold (being over 0.15, as described). This procedure was repeated until all the variables that had not been added to the model were above the p-value threshold and thus could not enter the model. Based on the stepwise linear regression and using the entire (discovery+validation) cohort (n=2876), the variables age, presence of antihypertensive treatment, study cohort, sex, BMI, presence of diabetes and eGFR remained in the final model and thus were considered for the SBP standardization. Similarly, for the DBP that was the case for the variables: BMI, sex, study, presence of antihypertensive treatment and age (but not eGFR or presence of diabetes). That said, for each patient, the BP measures were standardized, using each time the respective beta coefficients of the aforementioned, considered clinical variables, based on the formula:

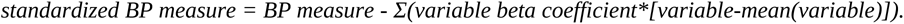

Last, using the entire discovery cohort (general population, n=749) a correlation analysis between these standardized BP measures (sSBP and sDBP), with the significant and consistently regulated peptides was also performed to reveal any relevant associations.

### Bioinformatics analysis

In an attempt to place the urinary proteomics findings in the biological context, bioinformatics analyses were employed. Protein-protein interactions were investigated using the STRING database, based on the default settings. The output was subjected to k-means clustering, based on the specified number of clusters (n=3).

### Software

The data processing and analyses were based on R programming^23^ (version 4.3.2) running on Ubuntu 22.04 computer software. The collection of R packages in the ‘tidyverse’ ^24^ package as well as the ‘broom’^25^ and ‘janitor’^26^ packages were extensively used. The random deviates were generated based on the runif function of the stats package, after applying the function set.seed(2020). Matching between hypertensive and normotensive participants at 1:1 ratio was based on the matchit function of the ‘MatchIt’ package ^27^. The Mann-Whitney U test for the peptide differential abundance comparison was performed based on the col_wilcoxon_twosample(exact=FALSE) of the ‘matrixTests’ package^28^. P-value<0.05 was the threshold for both the adjusted (for multiple testing) and the nominal significance. For the former, the Benjamini-Hochberg method was considered and performed via the function p.adjust(method=“BH”) of the ‘stats’ package. The stepwise linear regression was based on the function ols_step_both_p(pent=0.15, prem=0.15) of the ‘olsrr’ package^29^ using the output of the lm function of the ‘stats’ package with the clinical variables already described. The association of the peptides with the BP as a continuous variable was based on the rcorr(type=“spearman”) function of the ‘Hmisc’ package^30^.

Protein-protein interaction analysis and subsequent clustering, was based on STRING (https://version-12-0.string-db.org/)^31^. The linear regressions that considered the discovery and chronic disease fold changes were performed using ‘MedCalc’ software (version 12.1.0.0; MedCalc Software, Mariakerke, Belgium).

## Results

The entire discovery cohort included 749 individuals from the general population, while the validation cohort 2127 patients with type 2 diabetes. Based on the applied definition of hypertension, the discovery cohort consisted of 229 hypertensive and 177 normotensive, while the validation cohort consisted of 815 hypertensive and 106 normotensive individuals, reflecting the increased risk of hypertension in diabetic patients. Patients not being hypertensive (SBP≥140 mmHg and/or DBP≥90 mmHg), but with either SBP between 120 and 139 or DBP between 80 and 89 or taking antihypertensive medication were excluded from this analysis. The external matched cohort consisted of n=420 equally-numbered hypertensive and normotensive participants. The clinical and demographic characteristics of the discovery and the validation cohorts are given in **Table 1**. The study design and workflow are illustrated in **Figure 1**.

**Table 1.**
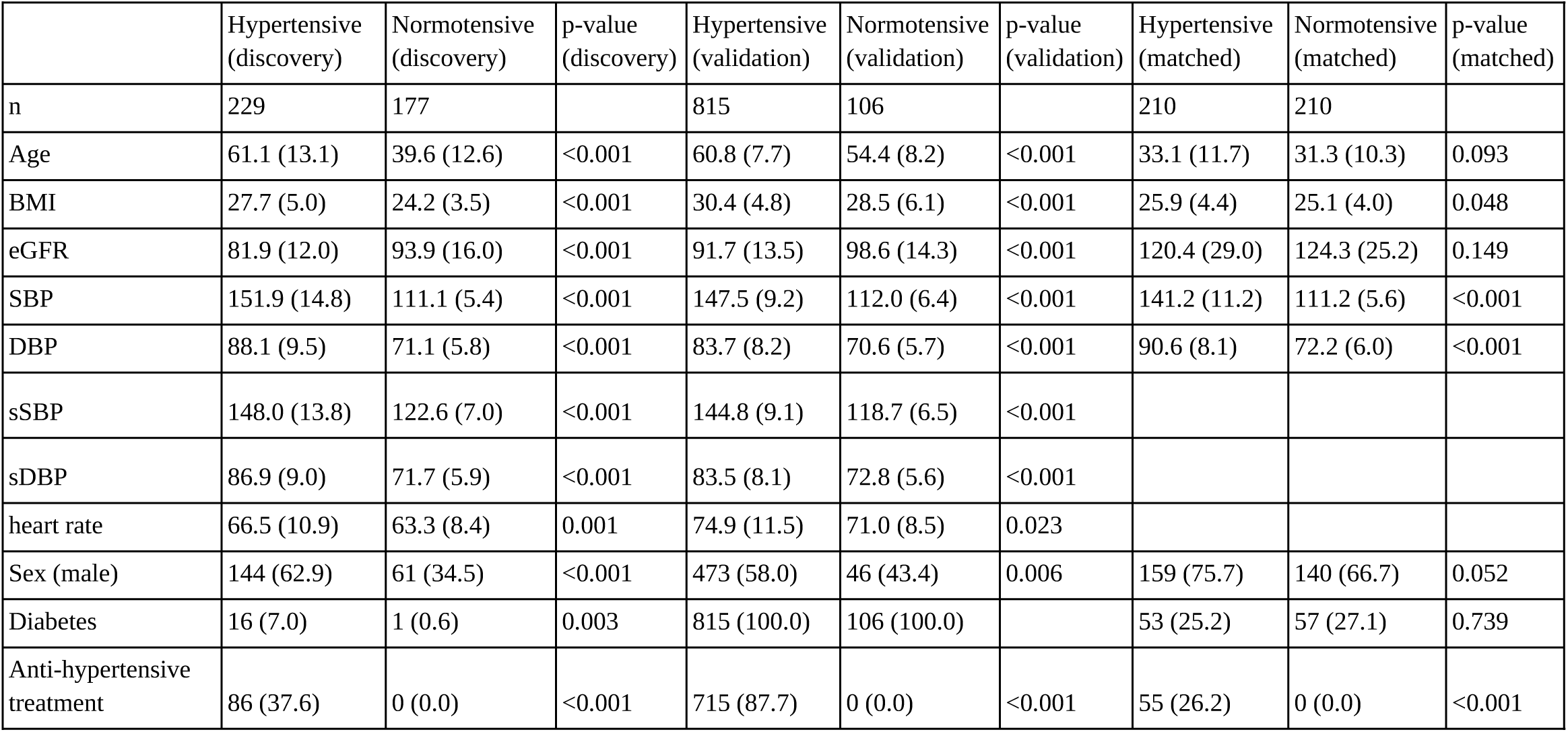
Clinical characteristics per cohort. Given is the number (n) of each entire group based on the BP values. A mean (standard deviation) or percentage (number) depending on the variable type is written for the available clinical information. BMI: body-mass index. eGFR: estimated glomerular filtration rate. sSBP: standardized systolic blood pressure. sDBP: standardized diastolic blood pressure.

### Urinary peptide associations with blood pressure

Using the discovery cohort, a comparison of the peptide levels between the hypertensive and normotensive groups was initially performed, revealing 308 peptides being significantly different (p-values adjusted for multiple testing). Of these, 205 were of the same abundance trend in both the discovery and validation cohorts, with 96 reaching nominal significance in the latter. As an additional step of evaluation, an extra peptide differential analysis was performed between the two groups, but this time using an external, matched cohort. The analysis confirmed a consistent regulation of 83 out of the aforementioned 96 peptides and only those were considered for subsequent analyses. These peptides were derived from 24 proteins, 12 of which were collagens (being the parental proteins of 68 peptides), namely COL1A1 (thirty-four peptides), COL3A1 (thirteen), COL2A1 and COL1A2 (four each), COL4A3 (three), COL16A1, COL18A1 and COL22A1 (two each) as well as COL13A1, COL15A1, COL21A1, COL25A1 (one each). As for the non-collagen proteins, three peptides were derived from FGA and two from MGP and one from ORM1, CD99, FGB, GSN, IGF2, KRT77, LTBP4, POTEF, PIGR and UMOD. The 20 most significant peptides (based on the discovery cohort) along with their regulation trends are listed in **Table 2**. To further confirm the relevance of these peptides with BP, their continuous correlation with sSBP and sDBP was investigated based on the Spearman’s rank method (using the entire discovery cohort of n=749). The range for the Spearman’s rank correlation coefficients for the sSBP and sDBP were -0.10 to +0.09 and -0.12 to +0.15, respectively. All the results and findings are listed in the **Supplementary table 1**.

**Table 2.**
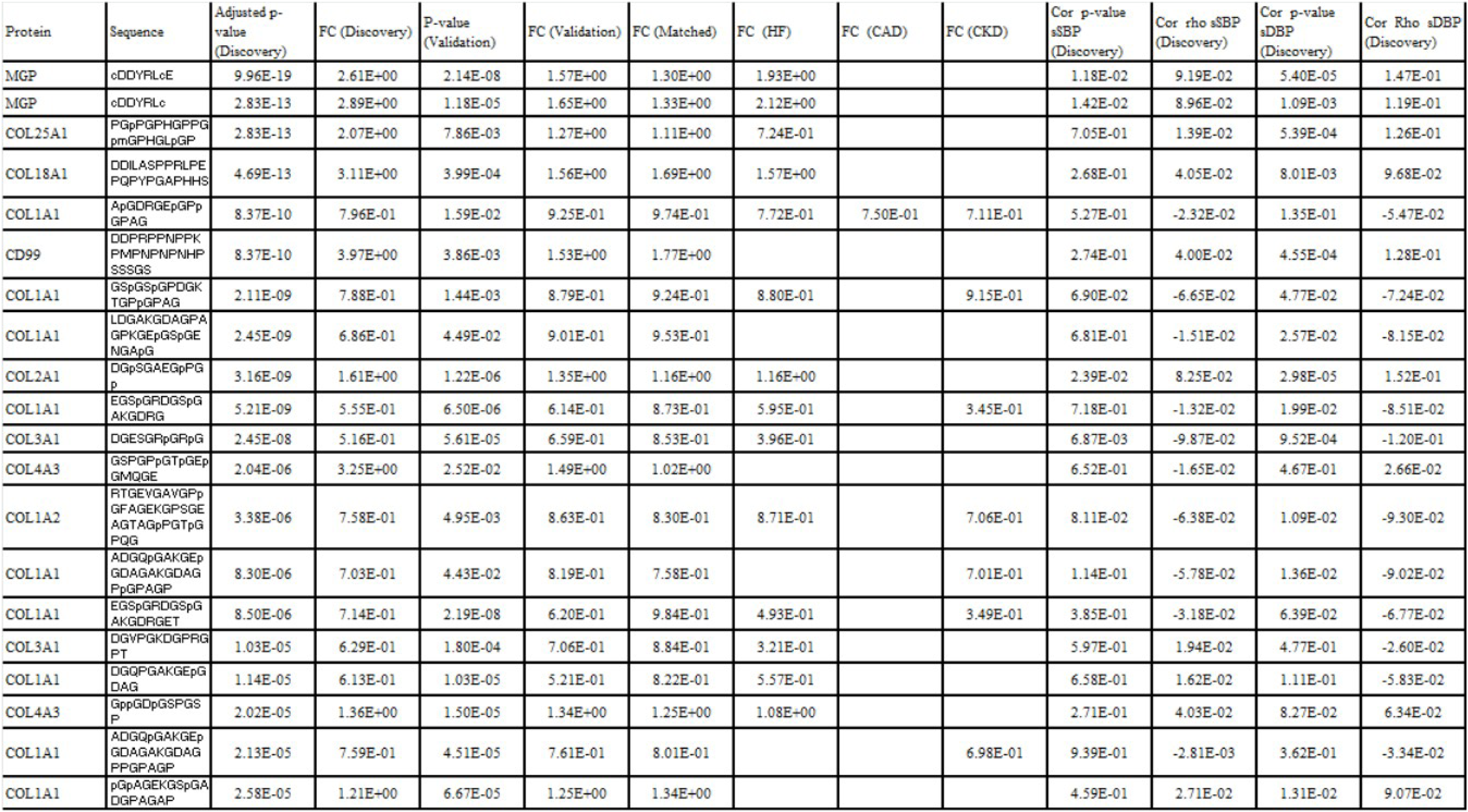
Results of the peptide differential abundance and Spearman’s rank correlation analyses as well as regulation of previously established biomarkers in chronic diseases. The analysis was performed separately for the discovery, validation and matched cohorts between hypertensive and normotensive groups. Listed are the 20 (out of the 83) most significant peptides identified as significantly different in both the discovery (p-value adjusted for multiple testing) and the validation cohorts (nominal significance) and demonstrating consistent regulation across both of these cohorts as well as the external matched cohort. Findings are sorted based on increasing adjusted p-values in the discovery cohort. Moreover, the fold change comparison with previously established biomarkers in chronic diseases revealed no substantial subclinical organ damage. Cor: correlation. HF: heart failure. CAD: coronary artery disease. CKD: chronic kidney disease. sSBP: standardized systolic blood pressure. sDBP: standardized diastolic blood pressure.

### Comparison with urinary peptides associated with kidney and cardiovascular disease

Although a cohort inclusion criterion was absence of clinical signs of end-organ damage, additional steps were performed with the aim to investigate the association of the defined 83 hypertension-associated peptides with previously established peptidomic biomarkers for chronic diseases, i.e., heart failure^8^, coronary artery^9^ and chronic kidney disease^10^. The results are listed in the **Supplementary table 1** for all 83 peptides. Of the 273 chronic kidney disease biomarkers, 25 (9.2%) peptides were common with the 83 hypertension-associated peptides, demonstrating significantly associated fold changes (R^2^=0.67, P<0.0001). In the case of the 577 urinary peptides that were described to be associated with heart failure, 44 (7.6%) peptides were common, with significant association in terms of regulation (R^2^=0.714, P<0.0001). Regarding the previously defined 160 coronary artery disease biomarkers, only 5 (3.1%) overlapped with our findings with regulation being, again, significantly associated (R^2^=0.96, P=0.0031). A further confounder with respect to subclinical end-organ damage may be proteinuria/albuminuria. However, none of these 83 peptides was derived from albumin or immunoglobulins, indicating that the observed changes do not appear to be connected to proteinuria.

### Bioinformatics analyses

In an attempt to gain insights into molecular mechanisms underlying the observed changes in the urinary proteome/peptidome, bioinformatics analyses were conducted investigating the 24 parental proteins of the 83 hypertension-associated peptides. The generated protein-protein interaction network consisted of 24 nodes and 84 edges, depicted in **Figure 2**. The protein-protein interaction enrichment yielded a significant p-value<1.0e^−16^. Within this network, a major cluster (mainly collagen-driven), comprising 14 nodes and 70 edges (PPI enrichment p-value< 1.0e^−16^), represented proteins involved in ECM remodeling. The second largest cluster (6 nodes, 6 edges, PPI enrichment p-value=2.89e^-07^), contained proteins involved in the innate immune system (i.e. FGA, FGB, ORM1, PIGR, GSN).

**Figure 2.**
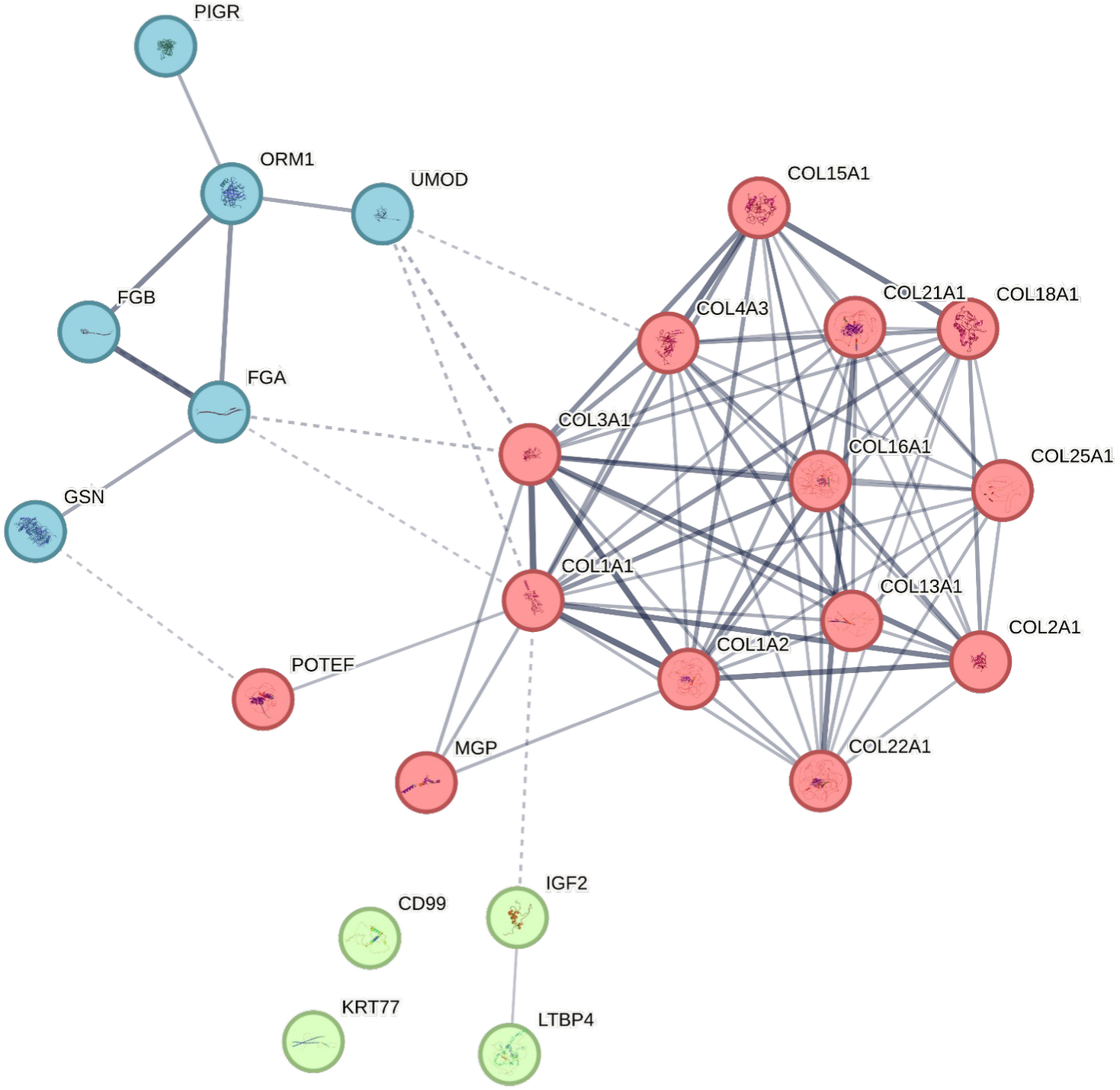
Protein-protein interaction network. The analysis was based on the 24 parental proteins of the 83 hypertension-associated peptides. The colors represent the three identified protein clusters using the STRING database. The red-colored cluster illustrates that predominantly the collagen-interplay appears to potentially be a large component of this condition.

## Discussion

Hypertension is both a consequence and a risk factor for numerous diseases. Given that there is a lack of proteomic studies into hypertension, we attempted to highlight the associated pathophysiology through the urinary peptidomics perspective. To this end, we performed case-control comparisons, using a large initial cohort of almost 3000 individuals without end-organ damage. Aiming towards high confidence into correct labelling of hypertensive and normotensive patients, we excluded patients not being hypertensive (SBP≥140 mmHg and/or DBP≥90 mmHg), but with either SBP between 120 and 139 or DBP between 80 and 89 or taking antihypertensive medication. Within a discovery-validation study design, we determined consistently regulated significant peptide changes between hypertensive and normotensive participants that were subsequently confirmed in an external cohort, matched for clinical confounders. In that way, we defined 83 hypertension-associated peptides of both collagen and non-collagen origin, which we then further investigated for correlations with the continuous standardized BP variables, and in protein-protein interaction bioinformatics analyses.

Although the molecular mechanisms of hypertension are complex and not completely known, several genome-wide association studies, as reviewed in^32^, have provided insights on genetic variants associated with BP, many of them mapping to non-protein-coding regions but also, in cases, colocalizing with quantitative trait loci (cis-eQTL) associated with renal expression of specific genes (e.g., Interleukin-1 receptor-associated kinase 1 binding protein 1 (IRAK1BP1), Microtubule-associated protein 1B(MAP1B), splice isoforms of NADH dehydrogenase (ubiquinone) complex I, assembly factor 6 (NDUFAF6), etc.). Additional features and molecular pathways associated to BP have been highlighted following integration of GWAS with kidney multi-omics include AGT (Angiotensinogen) and ACE (Angiotensin-converting enzyme), but also splicing isoforms of mitogen-activated protein kinase–associated protein 1 gene (MAPKAP1) and ubiquitin conjugating enzyme E2E 3 gene (UBE2E3), relevant to sodium reabsorption ^32^. Along the same lines, the uromodulin genetic locus has been strongly linked to hypertension, with carriers of a certain allele (SNP rs13333226) having both lower urinary uromodulin levels and lower hypertension risk^33,34^.

With regards to the relevant proteomic studies, one of the very few was conducted recently, by De Beer et al. in a population of young individuals ^35^. In that study, due to the low sample size, only a limited number of peptides remained significantly different between hypertensive and normotensive subjects after correcting for multiple-testing. Comparing the changes observed in that and in our work presented here, notable similarities were observed in groups from the general population, but not in diabetics. This likely reflects the fact that subjects included in the previous investigation were non-diabetics and of much younger age.

Since hypertension is a well-described risk factor for kidney and cardiovascular diseases, these patients, among others, were excluded from our study. However, this approach cannot rule out bias introduced due to subclinical organ damage. We therefore investigated if the identified hypertension-associated peptides were linked to kidney or cardiovascular disease in previous studies. Most of these peptides did not appear to be substantially associated with hypertension-mediated organ damage in chronic diseases. However, some similarities could still be noted. This overlap between peptides associated with hypertension and the previously defined peptide biomarkers for coronary artery disease, heart failure and chronic kidney disease might be reflective of shared disease pathophysiology patterns, revolving mainly around chronic inflammation and fibrosis. Consequently, the results indicate that urinary peptides may potentially provide information on chronic disease earlier than clinical parameters, before organ damage is established.

From the hypertension-associated peptides that were identified in our study, collagen-derived peptides were the majority. Collagens are among the most abundant proteins in the extracellular matrix (ECM), providing mechanical support to the vascular wall. Among major vascular diseases, ECM remodeling is a hallmark of hypertension, mainly occurring by collagenases or metalloproteinases (MMPs)^36^. Thus, it is not surprising that collagen fragments are very prominently associated with hypertension. The distribution and contents of ECM proteins varies with the type of vascular wall and its properties^37^. Fibrillar collagen types I and III comprise a notable portion of the vessel collagen localized in intima, media and adventitia ^37^ (e.g., in the aorta, type III dominates in the medial layer, while type I is more represented in the adventitia ^36^). The ratio of these two collagen types affects the mechanical properties of the arterial wall, with an increase in the proportion of collagen type I accounting for the ECM stiffening observed with aging^38^. That said, most of the hypertension-associated peptides that we defined in this study originated from these proteins. The majority of these peptides were found reduced in the urine of the hypertensive group, suggesting attenuated collagen degradation, consequently increased collagen deposits in these individuals. In addition to fragments of collagen I and III, differences between the two groups were also documented for the network-forming collagen type IV (COL4A3), potentially as a result of changes related to the basement membrane ^37^. Several processes may occur during hypertensive arterial stiffness as in a vicious cycle^36^. Collagen accumulation potentially leads to increased MMP activity (among other via increased protease expression as well as decreased expression of tissue inhibitors of metalloproteinases) to initiate ECM remodeling. Consequently, a context unfavorable for the stability of the relevant structures is generated. As a result, endothelial structure may be disorganized, resulting in infiltration of macrophages and mononuclear cells. This, in combination with the increasing presence of senescent cells in the vasculature due to aging-related processes, contributes to chronic inflammation. Further, as a compensatory mechanism an increase in collagen synthesis is observed, leading to the formation of disorganized, cross-linked accumulated collagens further contributing to vascular stiffness.

The molecular changes observed at the level of urinary peptides go beyond collagens. Matrix Gla protein is a vascular calcification inhibitor, activated by vitamin K through γ-glutamate carboxylation and serine phosphorylation^39^. The protein is highly expressed in the epithelium of the Bowman’s capsule and proximal tubules^40^, and also secreted by vascular smooth muscle cells and chondrocytes in the arterial tunica media ^41^, suggesting a role in vascular structures. Along these lines, increased plasma levels of inactive MGP forms were found positively associated with arterial stiffness and various forms of cardiovascular disease^42–46^, predictive of cardiovascular mortality^47^ and also negatively associated with eGFR^48^. We identified consistently increased urinary abundance of the two MGP peptides in the hypertensive group, which may be the result of increased degradation, hence reduced abundance in the body, which may result in attenuated protection from vascular calcification.

In the hypertensive patients a significant increase of a UMOD peptide was observed. UMOD is the most abundant protein in urine, secreted by the epithelial cells lining the thick ascending limb of Henle’s loop, modulating the sodium-potassium-two-chloride transporter^49^. In line with our findings and as mentioned above, a causal relationship between hypertension and UMOD levels has been suggested based on Mendelian randomization studies^50^.

Along these lines, in a recent manuscript urinary peptides were shown to predict the response to blood pressure medication^51^. Using the identified peptide changes along with the predicted impact of intervention on urinary peptides^52^ might help towards personalizing intervention and ultimately selecting the most appropriate therapy.

Our study has several limitations. Based on data availability, a case-control design had to be implemented. In addition, BP measurement was based on unstandardized methodology, which may have contributed to the observed weak peptide associations with continuous BP. Further, the study is based on previously collected datasets that were generated within different scientific designs.

## Conclusions

Our findings provide insights into the molecular alterations underlying hypertension. Deregulation in collagen type I and III peptides was associated with hypertension, likely indicating vascular ECM remodeling. Differences in the abundance of collagen peptides related to basement membrane were also observed. Finally, peptides from proteins involved in vascular calcification, sodium homeostasis, and sodium-potassium transport showed significant differences as well. The weak, but evident and significant association of peptides with BP appears biologically relevant towards highlighting the condition’s underlying mechanisms.

## Perspectives

Our findings confirm that changes in the vessel wall but also in tubular sodium transport and other subtle mechanisms occur very early in the natural history of hypertension, and more importantly that they are mirrored by changes in urinary peptides. The corresponding proteomic profile is largely different from that observed in patients with established kidney and/or cardiovascular diseases. While, in view of the multifactorial nature of hypertension, urinary proteomics is unlikely to become a diagnostic tool for hypertension, it may shed new light on early mechanisms of hypertension development. The changes observed in urinary peptides, particularly those derived from various collagen subtypes, point towards early vascular changes and thus provide insight into pathophysiology and may potentially direct the use of existing treatments or help developing new ones. These peptide changes may also deserve to be correlated with tissue and virtual histology of vessel wall, arterial stiffness and other biomechanical properties of the arteries.

## Data Availability

Data will be made available upon request directed to the corresponding author. Proposals will be reviewed and approved by the investigators and collaborators based on scientific merit. After approval of a proposal, data will be shared through a secure online platform after signing the data access and confidentiality agreement.

## Sources of Funding

This work was supported in part by the European Union’s Horizon 2020 research and innovation programme (860329 Marie-Curie ITN “STRATEGY-CKD”). Generation Scotland received core support from the Chief Scientist Office of the Scottish Government Health Directorates [CZD/16/6] and the Scottish Funding Council [HR03006] and is currently supported by the Wellcome Trust [216767/Z/19/Z]. The study was supported via the following funding: The Non-Profit Research Association “Alliance for the Promotion of Preventive Medicine” (URL: www.appremed.org) received a non-binding grant from OMRON Healthcare, Inc. Ltd., Kyoto, Japan.

## Disclosures

H.M. is the founder and co-owner of Mosaiques Diagnostics (Hannover, Germany). E.M., J.S., and A.L. are employed by Mosaiques Diagnostics. P.R. has received grants from Astra Zeneca, Bayer, and Novo Nordisk; and honoraria (to Steno Diabetes Center Copenhagen) from Astra Zeneca, Abbott, Bayer, Boehringer Ingelheim, Eli Lilly, Novo Nordisk, Gilead, and Sanofi.

## Supplemental Material

Supplementary table 1 (Mann-Whitney U test and Spearman’s rank correlation analysis results and known regulation in chronic diseases, for the 83 hypertension-associated peptides).

## Abbreviations

(BH): Benjamini-Hochberg
(BMI): body-mass index
(eGFR): estimated glomerular filtration rate
(BP): blood pressure
(SBP): systolic blood pressure
(DBP): diastolic blood pressure
(sSBP): standardized systolic blood pressure
(sDBP): standardized diastolic blood pressure
(ORM1): Alpha-1-acid glycoprotein 1
(CD99): CD99 antigen
(COL1A1): Collagen alpha-1(I) chain
(COL2A1): Collagen alpha-1(II) chain
(COL3A1): Collagen alpha-1(III) chain
(COL13A1): Collagen alpha-1(XIII) chain
(COL15A1): Collagen alpha-1(XV) chain
(COL16A1): Collagen alpha-1(XVI) chain
(COL18A1): Collagen alpha-1(XVIII) chain
(COL21A1): Collagen alpha-1(XXI) chain
(COL22A1): Collagen alpha-1(XXII) chain
(COL25A1): Collagen alpha-1(XXV) chain
(COL1A2): Collagen alpha-2(I) chain
(COL4A3): Collagen alpha-3(IV) chain
(FGA): Fibrinogen alpha chain
(FGB): Fibrinogen beta chain
(GSN): Gelsolin
(IGF2): Insulin-like growth factor II
(KRT77): Keratin, type II cytoskeletal 1b
(LTBP4): Latent-transforming growth factor beta-binding protein 4
(MGP): Matrix Gla protein
(POTEF): POTE ankyrin domain family member F
(UMOD): Polymeric immunoglobulin receptor (PIGR) Uromodulin

## Notes

### Author Declarations

This study is based on published, publicly available and anonymized data and thus does not require further IRB approval as stated by the ethics committee of the Hannover Medical School, Germany, under the reference number: 3116-2016.

